# Elevated blood pressure accelerates white matter brain aging among late middle-aged women: a Mendelian Randomization study in the UK Biobank

**DOI:** 10.1101/2023.04.06.23288211

**Authors:** Li Feng, Zhenyao Ye, Chen Mo, Jingtao Wang, Song Liu, Si Gao, Hongjie Ke, Travis A Canida, Yezhi Pan, Kathryn S Hatch, Yizhou Ma, Chixiang Chen, Braxton D. Mitchell, L.Elliot Hong, Peter Kochunov, Shuo Chen, Tianzhou Ma

## Abstract

**Background:** Elevated blood pressure (BP) is a modifiable risk factor associated with cognitive impairment and cerebrovascular diseases. However, the causal effect of BP on white matter (WM) brain aging remains unclear.

**Methods:** In this study, we focused on N=219,968 non-pregnant, family-unrelated individuals of European ancestry who had genotype data and two non-null clinical BP measurements available (99,532 male and 120,436 female, mean age=56.55, including 16,901 participants with neuroimaging data available) collected from UK Biobank (UKB). We adopted a chronological age-adjusted brain age metric, Brain Age Gap (BAG), as the outcome variable to measure the brain aging status. As a first step, we established a machine learning model to compute BAG based on white matter microstructure integrity measured by fractional anisotropy (FA) derived from diffusion tensor imaging data in a training set of subjects without hypertension (N=7,728). We then performed a two-sample Mendelian Randomization (MR) analysis to estimate the causal effect of BP on WM BAG in the whole population and subgroups stratified by gender and age brackets using two non-overlapping data sets (N=20,3067 for the set with genotype and BP data but no FA data; and N=8,822 for the set with genotype, BP and FA data). The main MR method used was generalized inverse variance weighted (gen-IVW) with other MR methods also included as sensitivity analysis.

**Results:** The hypertension group is on average 0.3098 years (95%CI=0.1313,0.4884; p <0.0001) older in WM brain age than the non-hypertension group of the same chronological age. Females are on average 0.8143 years (95% CI=0.6797 to 0.949; p <0.0001) younger in WM brain age than males of the same chronological age. The MR analyses showed an overall significant positive causal effect of diastolic blood pressure (DBP) on WM BAG, where every 10 mm Hg increase in DBP can lead to 0.371 years increase in brain age (CI: 0.034-0.709, p=0.0311). The stratified analysis by age and gender group found such significant causal effect of DBP on BAG to be most prominent among female women aged 50-59 (0.686 years/10mm Hg, CI: 0.054-1.318, p=0.0335) and aged 60-69 (0.962 years/10mm Hg, CI: 0.209-1.714, p=0.0122).

**Conclusion:** Hypertension and genetic predisposition to higher BP can accelerate WM brain aging specifically targeting at late middle-aged women, providing insights on planning effective control of BP for women in this age group.

## 1 Introduction

Elevated blood pressure (BP) is a primary risk factor for cerebrovascular and cardiovascular diseases [1, 2] and associated with a higher risk of cognitive impairment and dementia [3-5]. Studies have reported that high BP strongly correlates with alterations of both brain structure and neurobiological functions[6-8]. For example, a strong association was found between BP and cerebral white matter (WM) integrity measured by fractional anisotropy (FA) of water diffusion through diffusion tensor imaging (DTI) [4,5]. The microstructure of the human brain is constantly changing with normal aging, reflecting brain shrinkage, and cognitive and memory decline [9]. Thus, it is important to know how increased BP causes accelerated brain aging to reveal the underlying mechanism of BP on the brain and cognitive dysfunction. The prevalence of hypertension increases with age and differs among males and females in different age cycles [10-13], especially for older women after menopause. In this study, we will evaluate the causal effect of BP on WM brain aging in the general population as well as gender and age-specific groups to understand the benefits of controlling BP in reducing accelerated brain aging.

Brain age predicts chronological age from structural or functional neuroimaging features using machine learning (ML) algorithm [14-16] and has been commonly used to measure brain aging status. In our study, we will use multiple FA tract measures from DTI data to predict an age-adjusted white matter brain age metric, Brain Age Gap (BAG), which represents the difference between individuals’ brain age and their chronological age and regard it as the main outcome. Increases in BAG demonstrate evidence of accelerated aging and poorer brain health [17, 18]. The scalar BAG measure has been widely used in the literature as an alternative to multivariate imaging models to yield more robust and interpretable results in brain aging studies [19-22].

BAG can be influenced by multiple genetic, biological, environmental, and lifestyle factors [23]. Previous studies on BAG associated with lifestyle risk factors have focused on smoking and alcohol consumption [22, 24]. For high blood pressure and brain age, one study showed that elevated blood pressure was associated with brain aging, but no direct causality was reported [25]. In traditional observational studies, it is challenging to identify the causal relationship solely between high blood pressure and BAG, as other health risk factors related to abnormal brain aging, such as smoking, alcohol use, and chronic diseases, often co-occur with high blood pressure [26-29]. The Mendelian randomization (MR) framework has provided an unprecedented chance to overcome this challenge by using the genetic variants related to a risk factor of interest as a proxy for the risk factor to estimate its causal effect on the outcome, which is less prone to reverse causation and confounding [30-32]. The genetic determinants of BP are increasingly characterized in large-scale GWAS in the literature [10,15,16], which made it possible to identify better instruments for MR study. The MR methods have been successfully applied to investigate the causal effects of high blood pressure on myocardial infarction, atrial fibrillation, and other cardiovascular diseases as well as other chronic diseases [33-36], but no MR studies have looked at the causal effect of high BP on white matter brain aging in a large population or checked any different causal effects in age-or sex-stratifications.

To fill the gap, we used FA measures of DTI data from the UK Biobank (UKB) cohort to build an ML model to estimate WM BAG and performed a two-sample MR analysis to investigate the causal effect of BP (i.e., systolic blood pressure (SBP) and diastolic blood pressure (DBP)) on BAG using genetic variants associated with BP as instrumental variables (IVs) in the general population as well as stratified by gender and age groups. We hypothesized that 1) elevated BP would accelerate brain aging; 2) such causal effect is more prominent in more vulnerable age and gender groups. Our study highlights the age and sex disparities in the causal effects of modifiable cardiovascular disease-related risk factors on WM microstructure change, which provides implications for understanding the contributions of the late-life cognitive impairment risk.

## 2 Materials and Methods

### 2.1 Study Population and Variables

The data used in the present study were from the UK Biobank (UKB) cohort, which is a large-scale population-based study recruiting around 500,000 individuals aged 40 to 69 years and collecting comprehensive physical, genomic, health, and brain imaging phenotypic data [37]. We focused on non-pregnant, family-unrelated individuals of European ancestry (i.e., primarily British, and Irish) who had available genotypes, two non-null clinical BP measurements (SBP and DBP) from the initial assessment visits. We also excluded individuals with discordant reported versus genetically determined gender or whose DNA exhibited high rates of heterozygosity/missingness or have extreme values of white matter hyperintensities. The final pool includes N=228,473 individuals for analysis. To get a better genetically predicted BP, we further exclude those who have taken anti-hypertensive medicine [34]. The proportions of (non-)insulin-dependent diabetes mellitus, hypertensive heart disease, chronic ischemic heart disease, and other medical criteria are small, so we didn’t exclude those participants from our sample (Table 1 and Table S2). Figure 1 shows the number of subjects included at each step of the analysis.

**Table 1.** Characteristics of Participants for Association Analysis and Two-sample gen-IVW MR analysis in UK Biobank.

| Characteristics | Level | Non-HTN<br>Training Dataset | Genetic-Outcome Dataset |  | Genetic-Exposure Dataset |
| --- | --- | --- | --- | --- | --- |
|  |  |  | Non-HTN Testing<br>Dataset | HTN Testing<br>Dataset |  |
| n |  | 7728 | 7425 | 1397 | 203067 |
| Age (mean (SD)) |  | 54.23 (7.35) | 54.31 (7.36) | 58.48 (6.53) | 56.71 (8.02) |
| Sex (%) | female | 4215 ( 54.5) | 4116 ( 55.4) | 639 ( 45.7) | 111296 (54.81) |
|  | male | 3513 ( 45.5) | 3309 ( 44.6) | 758 ( 54.3) | 91771 (45.19) |
| SBP (mean (SD)) |  | 26.06 (3.96) | 26.07 (3.93) | 28.54 (4.64) | 137.97 (18.58) |
| DBP (mean (SD)) |  | 132.49 (16.48) | 132.38 (16.51) | 145.33 (17.95) | 82.25 (10.08) |
| BMI (mean (SD)) |  | 80.26 (9.38) | 80.01 (9.32) | 86.32 (10.64) | 27.23 (4.65) |
| Smoking Status (%) | Never | 4916 (63.71) | 4665 (62.83) | 797 (57.05) | 111814 (55.06) |
|  | Previous | 2351 (30.47) | 2332 (31.41) | 536 (38.37) | 70756 (34.84) |
|  | Current | 449 ( 5.82) | 428 ( 5.76) | 64 ( 4.58) | 20497 (10.09) |
| Alcohol drinker status (%) | Never | 159 ( 2.06) | 143 ( 1.93) | 28 ( 2.00) | 6036 ( 2.97) |
|  | Previous | 153 ( 1.98) | 122 ( 1.64) | 38 ( 2.72) | 6826 ( 3.36) |
|  | Current | 7414 (95.96) | 7160 (96.43) | 1331 (95.28) | 190205 (93.67) |
| Fruit Consumption (mean (SD)) |  | 3.07 (2.40) | 3.07 (2.28) | 3.13 (2.47) | 2.99 (2.48) |
| Vegetable Consumption (mean (SD)) |  | 4.72 (2.91) | 4.71 (2.71) | 4.94 (3.17) | 4.81 (3.08) |
| Sedentary Lifestyle (mean (SD)) |  | 4.16 (2.39) | 4.20 (2.33) | 4.80 (2.45) | 4.53 (2.48) |
| Insulin-dependent diabetes mellitus<br>(self-reported) (%) |  | 26 ( 0.34) | 14 ( 0.19) | 13 ( 0.93) | 1428 ( 0.70) |
| Non-insulin-dependent diabetes mellitus<br>(self-reported) (%) |  | 150 ( 1.94) | 119 ( 1.60) | 156 (11.17) | 10451 ( 5.15) |
| Hypertensive heart disease<br>(self-reported) (%) |  | 0 ( 0.00) | 0 ( 0.00) | 0 ( 0.00) | 194 ( 0.10) |
| Chronic ischaemic heart disease<br>(self-reported) (%) |  | 119 ( 1.54) | 90 ( 1.21) | 180 (12.88) | 12505 ( 6.16) |

**Figure 1.**
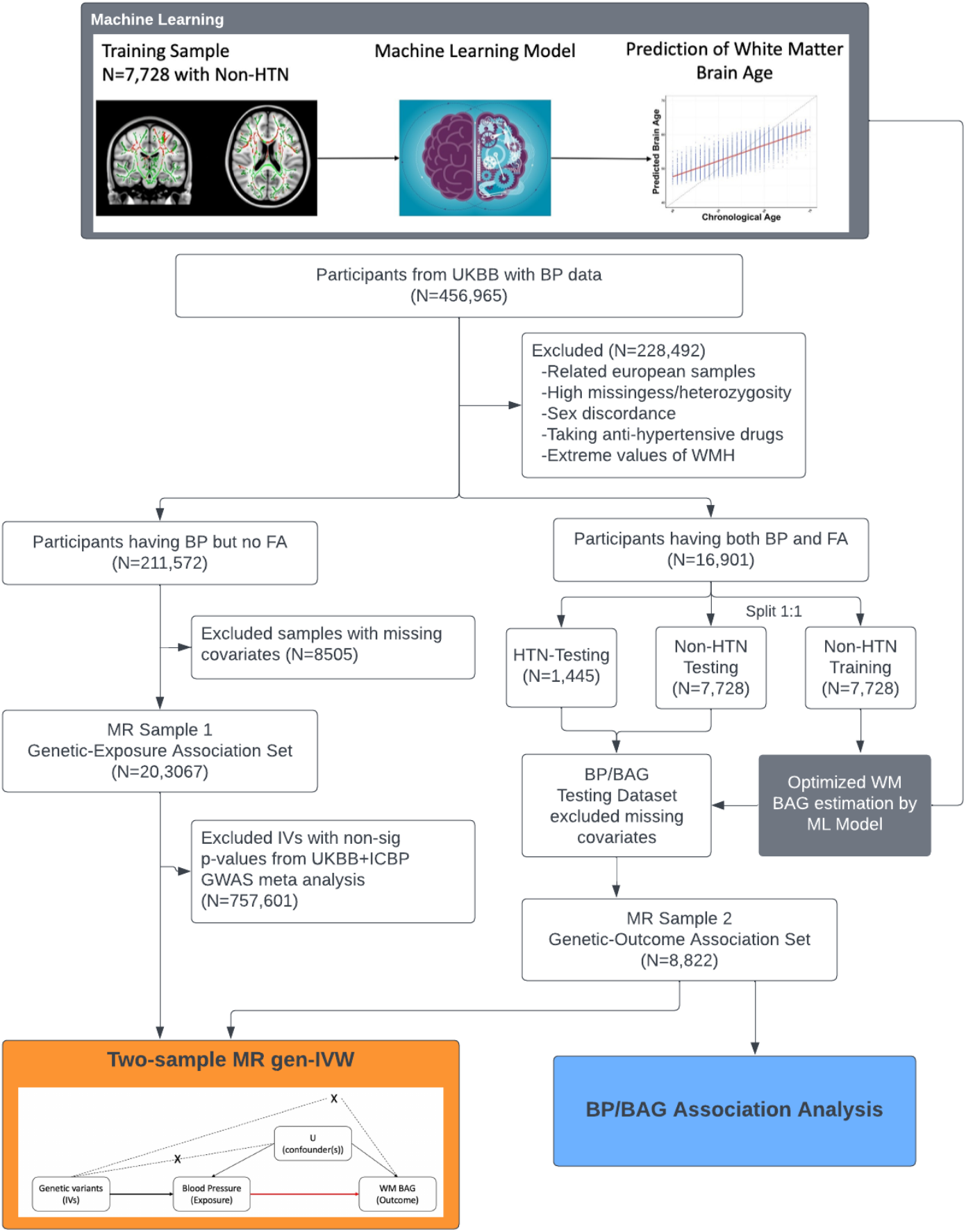
Flowchart of our main analysis procedures. Machine learning model was trained using training samples and tuned parameters with 5-fold cross-validation (CV) in a random forest regression (RF) model to determine an optimal locked model for the prediction of WM brain age. The locked model was applied to estimate WM brain age for testing samples and calculate WM BAG. Furthermore, 1) we corrected the bias of chronological age on WM BAG; 2) association analysis using general linear regression investigated the relationship between BAG and BP (i.e., SBP/DBP), controlling for related covariates; 3) we selected valid IVs by performing GWAS analysis on blood pressures (BP) (i.e., SBP/DBP) and checking IV assumptions. Finally, we conducted a gen-IVW analysis to investigate the causal effects of BP (i.e., SBP/DBP) on WM BAG using the previously selected valid IV.

#### Genotype data

The genotype data of participants provided by the UKB were assayed using two genotyping arrays, the UK BiLEVE Axiom Array and UK Biobank Axiom Array, which are described elsewhere [38, 39]. We further applied quality control for genotype data using the following criteria: minor allele frequency (MAF) ≥ 0.01, imputation quality score (INFO) > 0.1, Hardy-Weinberg equilibrium exact test p-value (HWE) ≥ 0.001, missing genotype rate (GENO) ≤ 0.05 and missingness per individual (MIND) ≤ 0.2.

#### Blood Pressure (BP) data

As the exposures of interest, we analyzed the following two BP traits in non-pregnant individuals with complete data collected from the initial (2006-2010) assessment visit: (1) Systolic blood pressure (SBP), calculated as the mean of two non-null BP measurements using phenotype codes 4080 in UKB; (2) Diastolic blood pressure (DBP), calculated as the mean of two non-null BP measurements using phenotype codes 4079 in UKB. Individuals who took antihypertensive treatments at baseline using phenotype code 20003 (Treatment/medication code) were excluded since their observed BP did not reflect the genetically predicted BP (see more details in the Supplementary Methods) [24]. Hypertension (HTN) was defined using International Classification of Diseases edition 10 (ICD-10) codes I10-I15 available in UKB. The ICD-10 codes I10-I15 represent the diagnosis of essential (primary) hypertension, hypertensive heart disease, hypertensive renal disease in hospital records or as a cause of death, or secondary hypertension, respectively [40, 41]. Hypertension status will be used to split the data for training and testing purposes.

#### Neuroimaging data

We used regional white matter (WM) Fractional Anisotropy (FA) measures collected from diffusion magnetic resonance imaging (dMRI) data from UKB imaging assessment starting from 2014 to compute the outcome of the study, the brain age gap (BAG). According to ENIGMA protocol, the per-tract mean value of each brain WM tract was calculated by using a Track-Based Spatial Statistics (TBSS) analysis from the DTI FA images [42]. FA measure, ranging from 0 to 1, represents the degree of anisotropy of a diffusion process and the integrity of cortical white matter [43]. A lower value of FA indicates less probability of diffusion in one direction (isotropic) [43]. This study focuses on FA data for a total of 39 tracts the covered multiple brain regions (see the list of 39 regional WM FA measures in Table S1). We excluded individuals with extreme values of the total volume of white matter hyperintensities (WMH) using phenotype code 25781 to minimize the bias [28].

#### Potential Confounders

We treated sex, age, BMI, alcohol consumption, smoking status, fruit and vegetable consumption, and sedentary lifestyle as potential confounders for our association analysis and MR analysis based on a previous study [44] (see more details in the Supplementary Materials). The continuous Age variable is categorized into 40-49, 50-59, 60-69 age groups. For both analyses, we conducted stratified analysis within each gender and age group.

#### Existing large-scale blood pressure GWAS summary data

To strengthen the IV selection step in two-sample MR analysis, in addition to UKB data alone, we also collected the largest existing GWAS summary data on BP from a meta-analysis of over 750,000 participants of European ancestry that combined a total of 78 different cohorts (mainly from International Consortium of Blood Pressure (ICBP) and part of UKB cohort) [16]. We used this existing meta-analyzed GWAS summary data to help select stronger and reproducible IVs in our two-sample MR analysis.

### 2.2 Overview of Analysis

Figure 1 shows a flowchart of our main analysis. In the first part, we will use machine learning (ML) technique to compute the outcome variable BAG among those participants with both BP and FA data available. We first split the data into a training set and a test set (1:1 split for participants with no hypertension). The training set only includes healthy subjects with no hypertension (Non-HTN Training; N=7728) to build an optimal ML model that computes BAG. The testing set includes independent groups of individuals without hypertension (Non-HTN Testing; N=7728) and individuals with hypertension (HTN Testing; N=1445) whose WM BAG can be estimated from the locked ML model obtained in the training set. In the second part, we will conduct a two-sample MR analysis to evaluate the causal effect on predicted BAG. The MR sample 1 Genetic-Exposure Association Set consists of participants with only BP but no FA data (N=203,067) to perform gene-exposure association analysis and select valid instrumental variables (IVs) in the two-sample MR analysis. To strengthen the IV selection step, we included the BP GWAS results from the meta-analyzed ICBP cohort (N=757,601; partially overlapped with UKB cohort) and only selected those IVs that passed the p-value thresholds (p<5 ×10^−8^) in both cohorts. The MR sample 2 Genetic-Outcome Association Set consists of non-overlapping participants in the testing set (Non-HTN + HTN testing excluding those with missing covariates; N=8822) who have both BP and BAG outcome variables available to perform gene-outcome association analysis in the two-sample MR analysis. [45]. The estimates from the two MR samples were combined using a generalized inverse variance weighted (gen-IVW) method to evaluate the causal effects of BP on BAG. In addition, we will also conduct an association analysis between BP and BAG in the Genetic-Outcome association set. For both MR analysis and association analysis, the analyses will be performed in the general population as well as stratified by gender and age groups.

#### Age Bias Corrected White Matter (WM) Brain Age Gap (BAG)

We used machine learning (ML) techniques to compute WM BAG based on 39 regional FA measures and the chronological age of individuals from the UKB. In the training set (non-HTN Training), we used random forest (RF) regression to generate a function for estimating unbiased brain age (Figure 1). The parameters of the RF regression were tuned based on the coefficients of determination (R^2^) between the chronological age and estimated brain age and mean absolute error (MAE) criteria to achieve the optimum predictive performance using a 5-fold cross-validation (CV). The RF regression was also used to select a set of FA features that have the most significant impact on brain aging. The locked ML model was then applied to the testing samples (i.e., non-HTN Testing and HTN testing datasets) to predict WM brain age. The WM BAG was calculated by subtracting individuals’ chronological age from their predicted brain age. The age-dependent bias has been noted to distort clinical interpretation in many brain age prediction studies [46-48]. We used a simple linear regression model [49] to remove brain age prediction bias from WM BAG and evaluated the performance of our correction method by the MAE.

#### BP/BAG Association Analysis

In BP/BAG association analysis, a general linear regression model was used to test the relation between BP and BAG in sample 2 Genetic-Outcome Association Set (N=8822), controlling the previously listed confounders, using Benjamini-Hochberg (BH) adjusted p-value < 0.05 [50]. In a sensitivity analysis, we redefined BP from two continuous variables (SBP/DBP) as one binary variable (see the definitions in supplementary material, Appendix 1. and the results in Table S7). To reduce the selection bias from the participants which excluded the group who took the anti-hypertensive medicine, we included the group of participants who took the hypertension medicine in the BP/BAG association model and provided the association result in Table S8. Also, we further adjusted menopause (Yes/No) and hormone replacement therapy (Yes/No) as covariates in the subgroup of the females aged 50-59 and the result was shown in Table S9.

#### Estimating causal effect by two-sample Mendelian Randomization (MR) analyses

In this study, we performed a two-sample MR analysis to evaluate the causal effects of BP on BAG treating candidate genetic variants as IVs. To implement the analysis (see details in supplementary material, Appendix 2.), we selected IVs based on BP GWAS (p<5 ×10^−8^) among MR sample 1 (N=203,067). We then performed a linkage disequilibrium (LD) clumping to remove genetic variants with r^2^>0.50 within a 1000-kb window) [51, 52]. Next, we removed IVs associated with any of the aforementioned confounders (Benjamini-Hochberg (BH) adjusted p-value > 0.05 [50]). To identify the strongest IVs and improve the power of MR analysis, we further excluded the IVs with a p-value greater than 5 ×10^−8^ from the large-scale existing BP GWAS summary data (meta-analyzed ICBP+UKB BP GWAS, N=757,601) [45]. Lastly, we performed conditional independence tests by regressing WM BAG on IVs given each BP to eliminate horizontal pleiotropic IVs (BH adjusted p-value > 0.1) using the MR sample 2 (N=8822).

We then applied the generalized inverse-variance weighted (gen-IVW) approach to estimate the causal effect of BP (i.e., SBP/DBP) on WM BAG using the previously selected valid IVs. The gen-IVW approach in MR analysis took the ratio of gene-outcome association and gene-exposure association estimates and combined multiple independent IVs into an overall estimate to assess the causal effect of exposure on the outcome while controlling the impact from LD between pairs of genetic variants [48, 53]. BH method was used to adjust for multiple comparisons in evaluating the causal effect of each BP on BAG. Because the impact of elevated BP on macrostructural WM degenerations can be modified by sex and age [54-56], in addition to analysis within the general population, we also performed age and sex stratified gen-IVW to explore the influence of chronological age and sex on causal discovery. We additionally applied sensitivity analyses using other popular MR methods including MR-PRESSO [57] for correcting horizontal pleiotropic outliers and MR-MIX [58] for adjusting genetic correlations. Besides, a leave-one-out approach [59] was used to evaluate the robustness of IVW results.

All statistical analyses were conducted using R (version 4.0.5) [60]. R packages, including “*MendelianRandomization*” (version 0.5.1) [61], “*MRPRESSO*” (version 0.1.0) [57], and “*MRMix*” (version 0.1.0) [58] were used to perform MR analyses.

## 3 Results

### 3.1 Descriptive statistics

We summarized the baseline characteristics of the ML training dataset and MR analysis datasets in Table 1. There is no systematic difference in the distribution of age, sex, BMI and BP between the non-HTN training group and non-HTN testing group (all p>0.05). The MR Genetic-Exposure Dataset consists of 91,771 males and 111,296 females, mean age of 56.71 (SD=8.02) years. The MR Genetic-Outcome dataset consists of 7425 participants (4116 males and 3309 females), mean age 54.31 (SD=7.36) in Non-HTN Testing Dataset and 1397 participants (639 males and 758 females), mean age 58.48 (SD=6.53) in HTN Testing Dataset. A very small proportion of individuals had self-reported diseases such as ((non-)insulin-dependent diabetes mellitus, hypertensive heart disease, and chronic ischaemic heart disease were also shown in Table 1 for all the groups. Table S2 presented a complete list of medical conditions among MR Genetic-Exposure Dataset and Genetic-Outcome Dataset.

### 3.2 Estimating White Matter Brain Age Gap

We used random forest regression to construct an ML model to predict BAG in the training set, locked the optimal model, and applied it to the testing set. The optimal model selected 25 FA measures for BAG estimation (Figures S1). After correcting age-bias on WM BAG, the predictive performance in testing data sets were obtained R^2^ of 0.94 (MAE=2.64 years, p < 2.2×10^−16^) for HTN test group and R^2^ of 0.94 (MAE= 2.48, p < 2.2×10^−16^) for non-HTN test group (Figure 2B). BAG was significantly different between HTN and non-HTN groups in all ages (p < 0.01) or different age strata (all p < 0.01) (Figure 2C-D). Furthermore, our data showed males had significantly higher WM BAG than females across all the age strata (all p < 0.01, Figure 2E). Overall, males had 0.8143 years (95%CI=0.6797 to 0.949; p <0.0001) higher than females regarding WM BAG. The detail of the age difference between males and females by age strata can be found in Table S4. Our results validate a previous finding that the adult female brain is on average a few years younger than the male brain in terms of brain metabolism [62].

**Figure 2.**
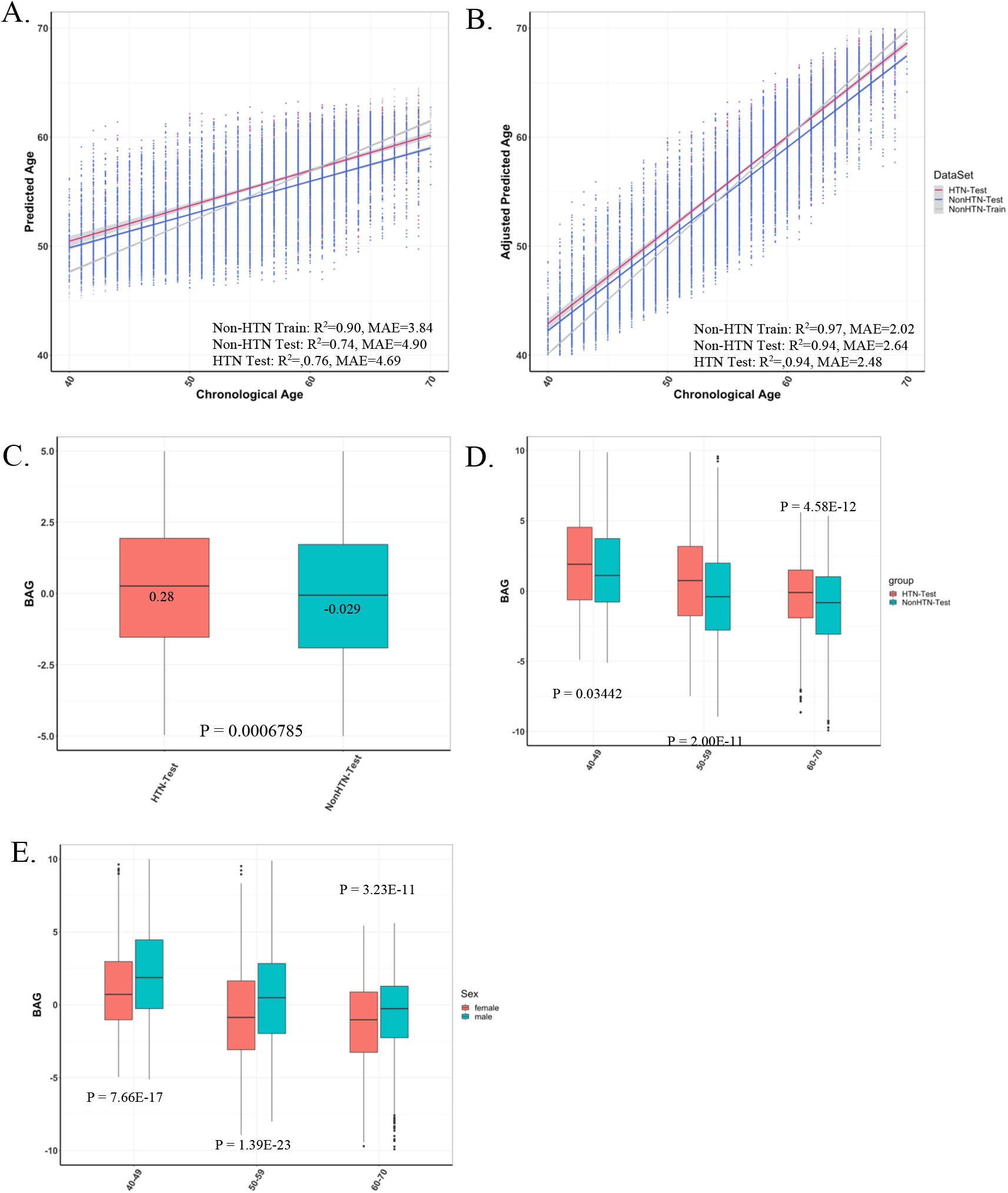
Linear scatter plots of WM BAG with/without correction for age (A, B) and box plots of WM BAG corrected by the bias of age between HTN and non-HTN from testing samples with/without age stratification (C, D). Box plots of age-adjusted WM BAG between females and males with age stratification (E).

**Figure 3.**
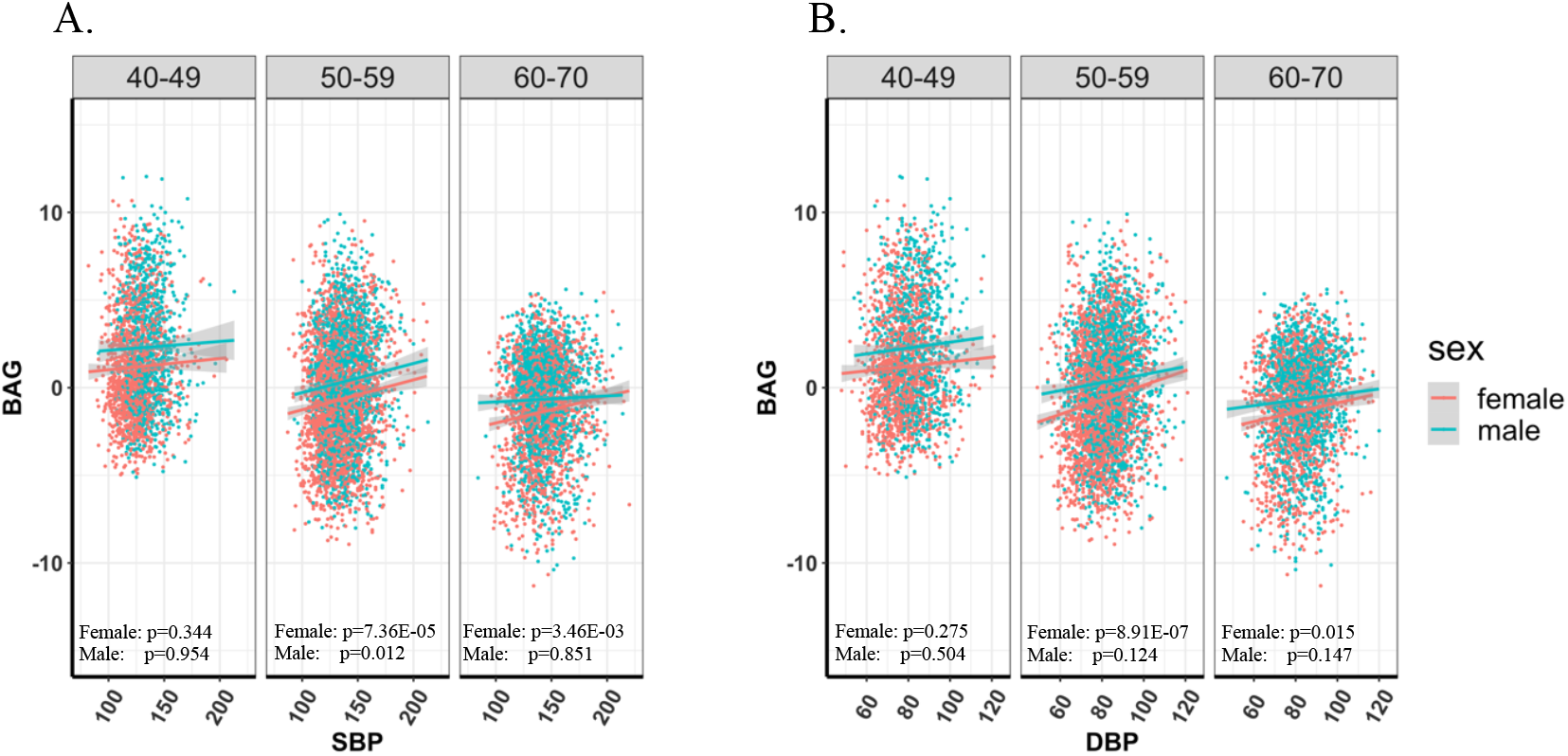
Scatterplots and linear fits illustrate relationships between BAG and BP (SBP and DBP) among different sex and age subgroup

### 3.3 Two-sample Mendelian randomization Analyses

GWAS analysis was conducted in the first gene-exposure association sample set. We identified 4813 and 5646 genetic variants with the p-value < 5 ×10^−8^ for SBP and DBP respectively. A majority of these single nucleotide polymorphisms (SNPs) were mapped to blood pressure-related genes (ex., *CACNB2, ATP2B1*, and *ARHGAP42*) that have been reported in previous studies [63-65] (full gene annotations were summarized in Table S5). After performing LD clumping (r^2^=0.5 within 1000-kb window) and removing IVs associated with the confounders (FDR>0.05), the results showed 187 and 195 genetic variants selected as potential IVs for SBP and DBP, respectively. After removing the IVs with a p-value greater than 5 ×10^−8^ from ICBP+UKB BP GWAS results, we got 180 and 191 SNPs for SBP and DBP, respectively, which indicates our IVs selection has high consistency with the selection from the largest existing GWAS results. Excluding the IVs the perform horizontal pleiotropy, the final numbers of valid IVs selected are 153 for both SBP and DBP, respectively. The numbers of IVs passing each step are summarized in Table S6.

We first tested associations between WM BAG and BP (i.e., SBP/DBP) controlling for the aforementioned confounders. Both SBP and DBP were positively associated with WM BAG (*β*_*SBP*_ = 0.0120, 95% CI = 0.0083 to 0.011, *p* = 4.30 ×10^−10^; *β*_*DBP*_ = 0.0175, 95% CI = 0.0111 to 0.0239, *p* = 1.31 ×10^−7^). With age and sex stratification, we found that both SBP and DBP were associated with WM BAG specifically in 50-59 (*β*_*SBP*_ = 0.0163, *p* = 3.74 ×10^−7^; *β*_*DBP*_ = 0.0259, *p* = 5.54 ×10^−6^) and 60-69 (*β*_*SBP*_ = 0.0086, *p* = 4.25 ×10^−3^; *β*_*DBP*_ = 0.0147, *p* = 7.11 ×10^−3^) age groups, and more significant association was found among female (*β*_*SBP*_ = 0.0149, *p* = 7.38 ×10^−9^; *β*_*DBP*_ = 0.0233, *p* = 3.59 ×10^−7^). The most significant association among the subgroups is for the females aged 50-59 (*β*_*SBP*_ = 0.0153, *p* = 2.45 ×10^−7^; *β*_*DBP*_ = 0.0352, *p* = 4.46 ×10^−6^) (Table 2). After applying the sensitivity analysis by including the participants who took anti-hypertensive medicine in the BP/BAG linear association analysis for all the groups or by further adjusting the menopause and hormone replacement therapy in the model in the females aged 50-59, the results of significance remained (Table S8 and Table S9).

**Table 2.**
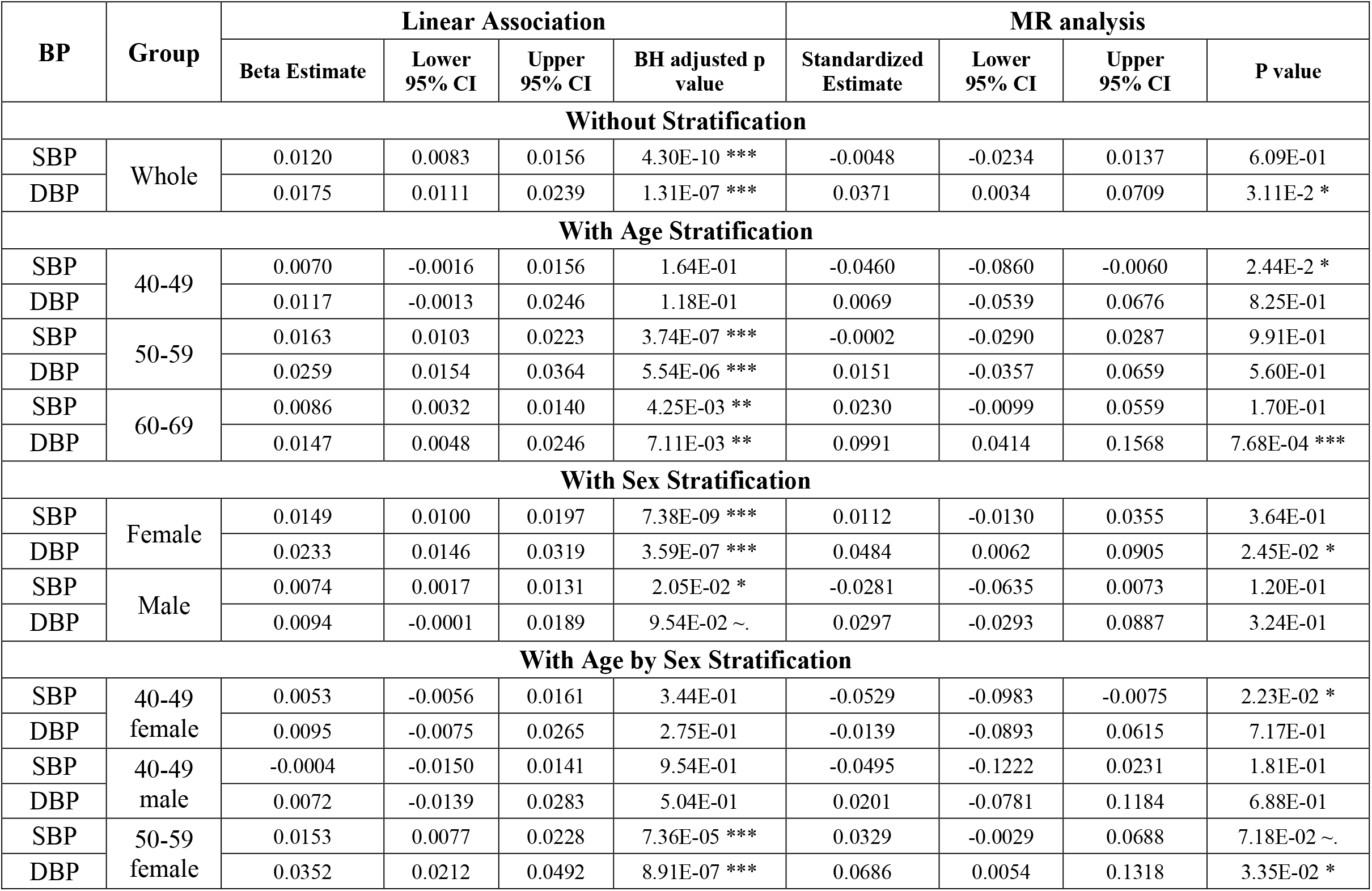
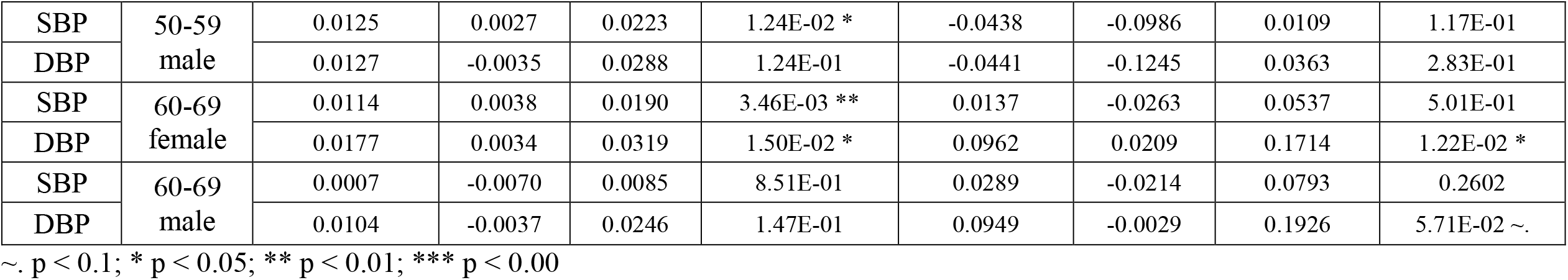
Results of linear association analysis between BP and WM BAG, and two-sample MR analysis (gen-IVW approach) between BP and corrected WM BAG adjusting for multiple potential confounders with/without stratifying age, sex, and age by sex.

The above results only showed association but no causal relationship due to the existence of reverse causality and confounding. Then we applied two-sample MR analyses to evaluate the causal effect of SBP and DBP on WM BAG. We found an overall significant causal effect of DBP (*β* = 0.0371, 95% CI = 0.0034 to 0.0709, *p* = 0.0311) but not SBP (p = 0.61) on WM BAG (Table 2), i.e. increasing 10 mmHg of DBP increases brain age by an additional 0.37 years. In stratified analysis, DBP had most significant causal effect on WM BAG in 60-69 age group (*β* = 0.0991, 95% CI = 0.0414 to 0.1568, *p* = 7.68 ×10^−4^) and among females (*β* = 0.0484, 95% CI = 0.0062 to 0.0905, *p* = 2.45 ×10^−2^). Late middle-aged females are most vulnerable to blood pressure change: Females in 50-59 age group (*β* = 0.0686, 95% CI=0.0054 to 0.1318, p = 0.0335) and 60-69 age group (*β* = 0.0962, 95% CI=0.0209 to 0.1714, p= 0.0122) had a significant causal effect for DBP on WM BAG, i.e. increasing 10 mmHg DBP increases brain age by additional 0.69-0.96 years in women aged 50-69. Those significant results were confirmed by implementing several sensitivity analyses using different MR methods and leave-one-out approach (see Table S7 and S8). On the other hand, a weak positive causal effect of DBP in age 60-69 (p=0.0571) was presented in the male group, and a weak causal effect for SBP in the 50-59 aged female group (p=0.0718). Some controversial results of the causal effects of DBP on WM BAG were shown among the male groups by reviewing all the MR analyses, which indicated that we could not draw any statistically reliable causal-effect conclusion for the male group. Find more details in Tables 2, S7, and S8.

## 4 Discussion

In this study, we used machine learning (ML) to calculate WM BAG. Further, we applied a linear model to correct the age-related bias on WM BAG regarding a healthy reference population without a diagnosis of hypertension (HTN). We found significant increases in WM BAG in HTN individuals and males compared to non-HTN individuals and females, respectively. The impact of HTN on brain WM integrity has been reported to be age-and-sex-dependent in previous studies [66-69]. Our overall association analysis between HTN and BAG confirmed that high blood pressure is highly related to elevated brain aging. Furthermore, we are the first to report that BP (i.e., SBP/DBP) has age- and sex-dependent causal effects on WM BAG implemented by the MR IVW approach, which is aligned with our stratified association analysis. In short, more significant causal effects of BP on WM BAG were observed among 50-to-69-age-group females.

To date, the patterns of brain age-related WM integrity decline are still unclear and controversial. For example, Voineskos et al., [70] identified WM aging patterns in cortico-cortical WM fiber tracts rather than interhemispheric association fibers. However, Zahr et al., [71] reported that compared to the posterior tracts, the effect of age on the reduction of FA was dominant at the anterior fiber tracts. A total of 25 selected FA features were obtained, involving both cortical-to-interhemispheric and anterior-to-posterior gradient distribution of WM tracts such as corpus callosum (GCC, BCC, and SCC) and bilateral anterior/posterior limb of the internal capsule (ALIC/PLIC), which might provide us with more comprehensive information for our brain-age prediction and further insights into the relationship of WM tracts with HTN in the model. Additionally, we observed individuals with HTN were likely to increase the aging process of WM integrity regardless of age group, which is consistent with previously reported findings [68, 72].

Previously, Christina S. Dintica el al. reported that time-weighted averages (TWA) SBP and DBP (per every 5-point mmHg) over the 30-year follow-up were associated with approximately 1 year and half a year greater brain age (MRI derived) respectively among 661 participants. Their results confirmed our findings but with some differences [73]. They reported a larger effect size in the association which may be due to the longer observation time, and they found SBP had a stronger effect than DBP which is opposite from our study. To improve the statistical power and investigate the inferring causality between BP and brain aging, we implemented two-sample MR approach in a large sample-size longitudinal cohort with high quality genotype, phenotype and brain imaging data. Our study is the first to demonstrate the causal discovery of BP (i.e., SBP/DBP) on WM BAG by combining ML and MR analysis from data in the large population UKB cohort. We revealed that genetic predisposition to higher BP (i.e., SBP/DBP) is associated with a greater risk of WM brain aging after overcoming confounding in observational studies. We found that DBP had a stronger causal effect on WM BAG than SBP. Additionally, many studies have revealed that the prevalence of HTN and brain aging increase with age and differs among males and females in different age cycles [10-13], especially for older adults and women after menopause. Overall, women have younger brains (neoteny) than males. Manu S. Goyal et al. [62] found that the adult female brain is a few years younger than the male brain over the entire adult life span in terms of brain metabolism, which is consistent with our findings regarding the sex difference in WM BAG across the year cycles (Figure 2E). On the other hand, interestingly, there is no association between BP and WM BAG among the age group of 40 to 49 for both genders, and a significant negative causal effect was detected among the 40-49 aged female group. Starting 50, the female group had much stronger causal effects of DBP compared to the male group. Also, the magnitude of causal effect of DBP on WM BAG in a female group was increased in older age group of 60 to 69 compared to that in the age group of 50 to 59, reflecting an age- and sex-dependent non-linear dose-response between BP and WM BAG that has also been reported in previous studies [74, 75].

Our study has found evidence showing a positive causal relationship between DBP and brain aging in late middle life, which is in line with other published studies [76-78]. On the other hand, naturally SBP tends to increase with age, but DBP may decrease with age due to changes in the cardiovascular system or other factors [79, 80]. The participates with higher SBP tends to have higher DBP across the middle-to-old-age strata [80]. Although people may reduce DBP in advanced age, a history of high blood pressure in midlife may have contributed to brain changes which may result in white matter lesions [78]. In addition, it’s also worth noting that lower DBP may confer risk (hypoperfusion). The relationship between blood pressure and brain aging is complex, multifactorial [81] or even non-linear [82]. Other factors, such as genetics, lifestyle, and comorbidities, may also contribute to brain changes and cognitive decline in advanced age. Further investigation to explore the multi-factors and non-linear relationships involved is warranted.

Our results showed that mid-to-older women with elevated blood pressure had a more causal effect on the risk of WM brain aging. This may be due to the gender difference in sex hormones and the vascular functions at molecular, cellular, and tissue levels [83]. Sex hormones produced among women of reproductive age may protect them from hypertension and related organ function (cardio vasculature, kidneys, and brain) [69, 84, 85]. Also, the animal and molecular studies revealed the sex differences in the mechanisms of BP control in linking renin– angiotensin–-aldosterone system (RAAS), sympathetic nervous system (SNS), immune system, nitric oxide, endothelin 1 (ET1), and vasopressin [83, 86-89]. Most studies studied the mechanism of BP related to the cardiovascular system and renal function. We provided novel insights on how BP causes increasing in brain aging in humans, especially in late middle-aged women, and affects differently in an age- and sex-dependent manner, but the causal effects warrant further investigations. For clinical practice, women after menopause should receive more attention on high blood pressure prevention or hypertension treatment to prevent brain aging and related brain diseases (Alzheimer’s Disease, cognitive decline, etc.).

The study has several strengths. First, the two-sample MR approach represents a robust method for inferring causality between a modifiable exposure and an outcome in observational studies using genetic variants as instrumental variables. We applied this method to two large cohorts, primarily the UKB cohort, and subsequently the UKB+ICBP cohort for validation, utilizing high-quality data, thereby enhancing statistical power, and facilitating a more comprehensive understanding of the causal association between blood pressure and WM BAG. This method allowed for the reduction of confounding factors and reverse causation, thus enhancing the validity of the causal inference. Second, we calculated WM BAG using ML and adjusted for age bias. Third, we used both linear association and a two-sample MR method to make an association test and a causal inference of BP on WM BAG in a large dataset with high-quality genotype and phenotype data. Fourth, we restricted our study population to family-unrelated individuals of European ancestry to remove the population stratification confounding effect. Fifth, we exclude participants taking antihypertensive drugs and the extreme value of FA. Lastly, we conducted several sensitivity analyses in both association tests and two-way MR to provide a more valid and robust estimation.

Our study has several limitations. First, our study only included the participants without taking anti-hypertensive medicine, which aims to include a better genetically predicted BP data, and hence the ascertainment bias may have been introduced. Reassuringly, we have done a sensitivity analysis in the BP/BAG association analysis to confirm our findings by including the participants who took anti-hypertensive medicine, suggesting that any such bias is not likely to be affecting our conclusions. Second, the reversal causality from brain aging on BP may require further investigation (i.e., bidirectional MR), although the reverse causality from our study is somewhat mitigated [90]. Third, due to the lack of brain-imaging data for younger individuals (i.e., age < 40), the potential impact of HTN on brain WM aging over time is not investigated in our current study. In addition, other potential factors associated with brain aging, such as smoking, alcohol intake, and depression, would be involved in further dedicated studies to provide more persuasive evidence for the prediction of brain age. Although we demonstrated significant causal effects of BP (i.e., SBP/DBP) on WM BAG, the molecular mechanisms of selected valid IVs remain unclear. It is critical to integrate other advanced methods (i.e., transcriptome-wide association analysis) and/or to use multiple omics data (i.e., proteins and RNA) that would allow us to obtain a better understanding of the biological system underlying the causal pathway between BP and WM BAG. Finally, although the individuals recruited in the UKB cohort were more likely to have healthier medical conditions and higher socioeconomic status, findings regarding exposure-disease relationships are still reliable and generalizable to other populations reported in previous study [91].

In conclusion, our study has shown that the non-hypertension group and females had younger brains (WM BAG) across all the mid-to-older year cycles. Hypertension and genetic predisposition to higher BP (i.e., SBP/DBP) can accelerate brain WM aging in an age- and sex-dependent manner. These findings shed insight on understanding the abnormal WM brain changes due to the modifiable risk factors of cardiovascular disease and further provide evidence to reduce the incidence of late-life cognitive impairment and dementia, reflecting the effective control of BP in age and gender awareness would be essential for rational planning of health services.

## Data Availability

All data produced in the present study are available upon reasonable request to the authors

https://www.ukbiobank.ac.uk/

## 6 Author Contributions

## 8 Acknowledgments

## 9 Data Availability Statement

The raw genetic and phenotypic data used for this study can be found in the UK Biobank (http://www.ukbiobank.ac.uk/)

