## Supplementary tables and figures for "Elevated blood pressure accelerates white matter brain aging among late middle-aged women: a Mendelian Randomization study in the UK Biobank": Supplementary Material.pdf

**Appendix 1. Definitions of BP as a binary variable in BP/BAG Association sensitivity analysis**

In a sensitivity analysis, the linear association analysis was applied to examine the high blood pressure as three different defined binary variables (diagnosed, stage-2 high blood pressure, and combined) and WM BAG adjusting for multiple potential confounders with/without stratifying age, sex, and age * sex. BP as binary variable 1: define high blood pressure as diagnosed only (International Classification of Diseases edition 10 (ICD-10) codes I10-I15 available in UKB). BP as binary variable 2: define high blood pressure as stage-2 high blood pressure SBP >139/DBP >89. BP as binary variable 3: define high blood pressure as a combined diagnosed and stage-2 high blood pressure.

**Appendix 2. Detailed IVs selection for the two-sample Mendelian randomization**

Following by the IVs selection assumptions [35], in the first data set, we restricted to the participants with both the genotype and BP data but no BAG data to investigate the gene-exposure associations (MR sample 1, N=203,067). We first performed genome-wide association study (GWAS) analyses on BP (i.e., SBP/DBP) in the Genetic-Exposure sample (Fig. 1B) to identify genetic variants strongly associated with BP (i.e., SBP/DBP) using a stringent genome-wide p-value threshold at $5 \times 10^{-8}$ under an additive genetic model adjusted for sex, age, body mass index (BMI), genotyping chip type and top 10 principal components (PCs) of population admixture. To identify the strongest IVs and improve the power of MR analysis, we then used the large-scale existing BP GWAS summary data (meta-analyzed ICBP+UKBB BP GWAS) to exclude the IVs with the p-value less than $5 \times 10^{-8}$ [31]. We further performed a linkage disequilibrium (LD) clumping (with $r^2$>0.50 within 1000-kb window) [36, 37] to confine IVs independently using PLINK (version 1.9, www.cog-genomics.org/plink/1.9/[38]). Next, we removed IVs associated with aforementioned confounders using Benjamini-Hochberg (BH) adjusted p-value > 0.05 [39], which indicates no association. In the second set, we included the participants with all the genotype, BP, and BAG data to investigate the gene-outcome associations (MR sample 2, N=8822) and the outcome-exposure associations (BP/BAG association analysis). There were no shared individuals between these two sets of samples. Lastly, we performed conditional independence tests by regressing WM BAG on IVs given each BP to eliminate horizontal pleiotropic IVs (BH adjusted p-value > 0.1) using the Genetic-Outcome sample (Fig. 1B). Additionally, we followed an MR guidance for IV selections proposed by Burgess et al., [40] to confirm the remaining IVs with a biological link to BP (i.e., SBP/DBP) listed in the NIH National Human Genome Research Institute GWAS catalogue (http://www.genome.gov/gwastudies) and annotate them from FAVOR (http://favor.genohub.org/) [41]. These criteria substantially enhance the creditability of causal role of risk factor on outcome.
